# Improved precision oncology question-answering using agentic LLM

**DOI:** 10.1101/2024.09.20.24314076

**Authors:** Rangan Das, K Maheswari, Shaheen Siddiqui, Nikita Arora, Ankush Paul, Jeet Nanshi, Varun Udbalkar, Apoorva Sarvade, Harsha Chaturvedi, Tammy Shvartsman, Shet Masih, R Thippeswamy, Shekar Patil, S S Nirni, Brian Garsson, Sanghamitra Bandyopadhyay, Ujjwal Maulik, Mohammed Farooq, Debarka Sengupta

## Abstract

The clinical adoption of Large Language Models (LLMs) in biomedical research has been limited by concerns regarding the quality, accuracy, and reliability of their outputs, particularly in precision oncology, where clinical decision-making demands high precision. Current models, often based on fine-tuned foundational LLMs, are prone to issues such as hallucinations, incoherent reasoning, and loss of context. In this work, we present GeneSilico Copilot, an advanced agent-based architecture that transforms LLMs from simple response synthesizers to clinical reasoning systems. Our approach is centred around a bespoke ReAct agent that orchestrates a suite of specialized tools for asynchronous information retrieval and synthesis. These tools access curated document vector stores containing clinical treatment guidelines, genomic insights, drug information, clinical trials, and breast cancer-specific literature. To leverage large context windows of current LLMs, we implement a hybrid search strategy that prioritizes key information and dynamically integrates summarized content, reducing context fragmentation. Incorporating additional metadata further allows for precise, transparent and evidence-backed reasoning at each step of the thought process. The system ensures that at every stage, the agent can synthesize meaningful, context-aware observations that contribute to a coherent and comprehensive final response that aligns with clinical standards. Evaluations on real-world breast cancer cases show that GeneSilico Copilot significantly improves response accuracy and personalization. This system represents a critical advancement toward making LLMs clinically deployable in precision oncology and has potential applications in broader medical domains requiring complex, data-driven decision-making.

## INTRODUCTION

Cancer’s inherent complexity, driven by both inter and intra-tumoral heterogeneity, presents a significant hurdle in clinical management. This genetic heterogeneity allows tumors to evade traditional treatments. However, the advent of precision oncology has led to the development of genome-targeted and genome-informed therapies, which aim to address this challenge. From 2006 to 2020, the eligibility for genome-targeted therapies in the U.S. increased from 5.13% to 13.60%, while the response rate improved from 2.73% to 7.04%. Similarly, genome-informed therapies saw a rise in eligibility from 10.70% to 27.30% and an increase in response from 3.33% to 11.10% during the same period. Interestingly, most of the eligibility increase for genome-targeted therapies occurred after 2018, whereas most of the response increase was observed before 2018. These findings highlight a concerning trend: while eligibility for these therapies is on the rise, the actual response rate remains low^1^. Large Language Models (LLMs) offer a powerful opportunity to address these challenges^2^. By leveraging their ability to process and synthesize vast amounts of healthcare data, LLMs can assist oncologists in navigating the ever-expanding landscape of targeted therapies. They can analyze a patient’s specific genetic profile, identify relevant clinical trials and treatment guidelines, and even suggest potential drug combinations tailored to the unique characteristics of the patient’s cancer considering comorbidities and side effects. While LLMs can correctly identify some key strategies and offer reasonable, albeit incomplete, suggestions even experts missed, they can also generate factual errors (hallucination), irrelevant, harmful, or biased content^3^.

While multiple industries have adopted LLM-based models, adoption in biology and medicine is still lacking^4^. With the ever-evolving treatment guidelines and drug approval^5,6^ it is difficult for fine-tuned LLMs to stay updated. State of the art models like Med-PaLM ^7^, BioGPT ^8^, and BioBERT ^9^ are standalone systems that are trained and fine-tuned on domain-specific large-scale biomedical corpora. These have achieved notable results on medical question answering datasets. GatorTron^10^ is a similar model that has been designed and trained from scratch, and subsequently fine-tuned for tasks like clinical concept extraction, medical relation extraction, semantic textual similarity, natural language inference, and medical question answering. While there are domain-specific LLMs, they come with the same drawbacks of foundational models. Foundational models such as ChatGPT 3.5 itself can act as a support tool for breast tumor board, but suffer from the lack of references, or the potential to produce seemingly credible but incorrect responses^11^. In the domain of radiation oncology, ChatGPT 3.5 showed high accuracy and completeness in radiation oncology queries, but higher-than-recommended readability levels suggest the need for refinement for improved patient accessibility and understanding^12^. Newer models such as GPT-4 and models from Anthropic provide better responses, but all LLMs continue to have clinically significant error rates, including examples of overconfidence and consistent inaccuracies^13^. In the context of treatment guidelines, ChatGPT provides concise, accessible supportive care advice including many non-medical support recommendations, but its recommendations lacked the specificity observed in National Comprehensive Cancer Network (NCCN) guidelines including often not suggesting any medications^14^. These discrepancies with guidelines raise concerns for patient-facing symptom management recommendations. This can be partly attributed to the fact that these models are trained on publicly available data and do not have sufficient specialized domain information. Models implemented on more focused domains have better performance. CancerBERT^15^ is an example of a model trained on a narrower domain, but the model has only been evaluated on named entity recognition (NER) tasks. The output of standalone models can be further optimized through retrieval augmented generation (RAG). In RAGs, the LLM retrieves information from pre-defined storage and synthesizes the response. External data can be used to augment the response, thereby providing more context and reducing false information. For example, in clinical trial screening, GPT-4 has shown promising performance when augmented with external data sources^16^. Similarly, GPT-4 has also been used for retrieving cancer guidelines. GPT-4 with RAG provided significantly higher correct responses when compared to the standalone LLM service^17^. RefAI^18^ is a similar tool that uses retrieval augmented generation to fetch medical literature in real time and summarize them. These examples show the potential of retrieval augmented generation to mitigate the shortcomings of standalone LLM services.

While LLMs with extended context windows—such as those seen in models like Gemini 1.5—show improvements in retrieving relevant facts, these models still fall short of maximizing recall. For example, Gemini 1.5 demonstrated superior accuracy over GPT-4 at shorter context lengths but experienced a noticeable drop in recall when dealing with extended contexts^19^, achieving only around 60-80% recall at higher token limits. This gap means that even with the ability to handle massive amounts of information, the LLM may fail to retrieve a significant portion of available relevant data. This loss significantly undermines the system’s reliability in high-stakes fields like precision oncology, where critical pieces of information must not be overlooked.

While simple Retrieval-Augmented Generation (RAG) systems attempt to address the challenges of context and relevance by fetching documents to augment the LLM’s output, they often reduce the LLM to the role of a passive synthesizer. These systems rely heavily on external data sources, which the LLM incorporates without engaging its deeper reasoning capabilities. As a result, the LLM may act more like a writing assistant, handling content without participating in any meaningful decision-making process. This approach can be limiting, particularly in complex fields like precision oncology, where critical clinical decisions depend on integrating insights from multiple verticals—such as genomic profiles, treatment guidelines, and clinical trials. Simple retrieval mechanisms cannot adequately navigate these diverse data streams or provide the nuanced reasoning required for personalized patient care.

To overcome these limitations, it is essential to move towards agentic systems that not only retrieve information but also actively reason over it. Unlike traditional RAG systems, which relegate LLMs to performing static retrieval tasks, agent-based frameworks empower LLMs to function as dynamic reasoning systems. In these frameworks, the LLM is not just a passive receiver of information, but an active participant in the decision-making loop, integrating data from various sources and engaging in logical synthesis. The GeneSilico Copilot (GSCP) exemplifies this agentic approach. Powered by a bespoke ReAct agent ^20^, GSCP actively orchestrates multiple tools to retrieve and process information from curated sources. This multi-tool architecture allows the system to engage in complex reasoning, synthesizing insights from a hybrid retrieval mechanism while dynamically filtering out irrelevant data. By leveraging these tools, GSCP transcends the limitations of both long-context-window models and basic RAG systems, ensuring that the LLM generates transparent, evidence-based responses that are contextually aware and clinically relevant. This agentic reasoning system is vital in oncology, where accurate and context-specific responses can affect treatment choices.

## RESULTS

### GSCP, an agentic framework for precision oncology

Precision oncology requires the integration of detailed information from multiple domains, such as drug dosages, treatment guidelines, and clinical trials. A single retrieval system is often inadequate for navigating this complexity. To address this, we propose an agent-based approach that enables the LLM to dynamically control multiple specialized tools based on conversational context.

At the core of the GeneSilico Copilot (GSCP) is a modified ReAct agent, which improves upon the traditional reason-observation-action loop. When compared to commercially available language models services, what sets GSCP apart is its ability to show the thoughts and observations, along with links to the document sources while generating a response. The agent operates with two types of memory: conversational memory for user interactions and working memory for managing tool-based processes. This distinction ensures coherent communication with both users and tools. The observations from the tools may often serve as additional related information for oncologists. These include treatment guidelines, drug dosage information, adverse reactions and other such information.

The GSCP retrieval system integrates a vector store with hybrid search (implemented via Qdrant) and a language model. The vector store handles semantic retrieval, while the language model generates insights based on the entire user query. After retrieving documents, the system re-ranks them for relevance before processing them further to create context-aware insights.

While both the agent and tools utilize language models for interaction, only the agent makes decisions, driving the overall reasoning process. This process involves analysing the query, retrieving and filtering relevant information via tools, and synthesizing a comprehensive response. Tools play a crucial role in generating insights by filtering retrieved documents to those most relevant to the query. The agent then uses these insights to formulate a refined, evidence-based response (**Figure 1**).

**Figure 1:**
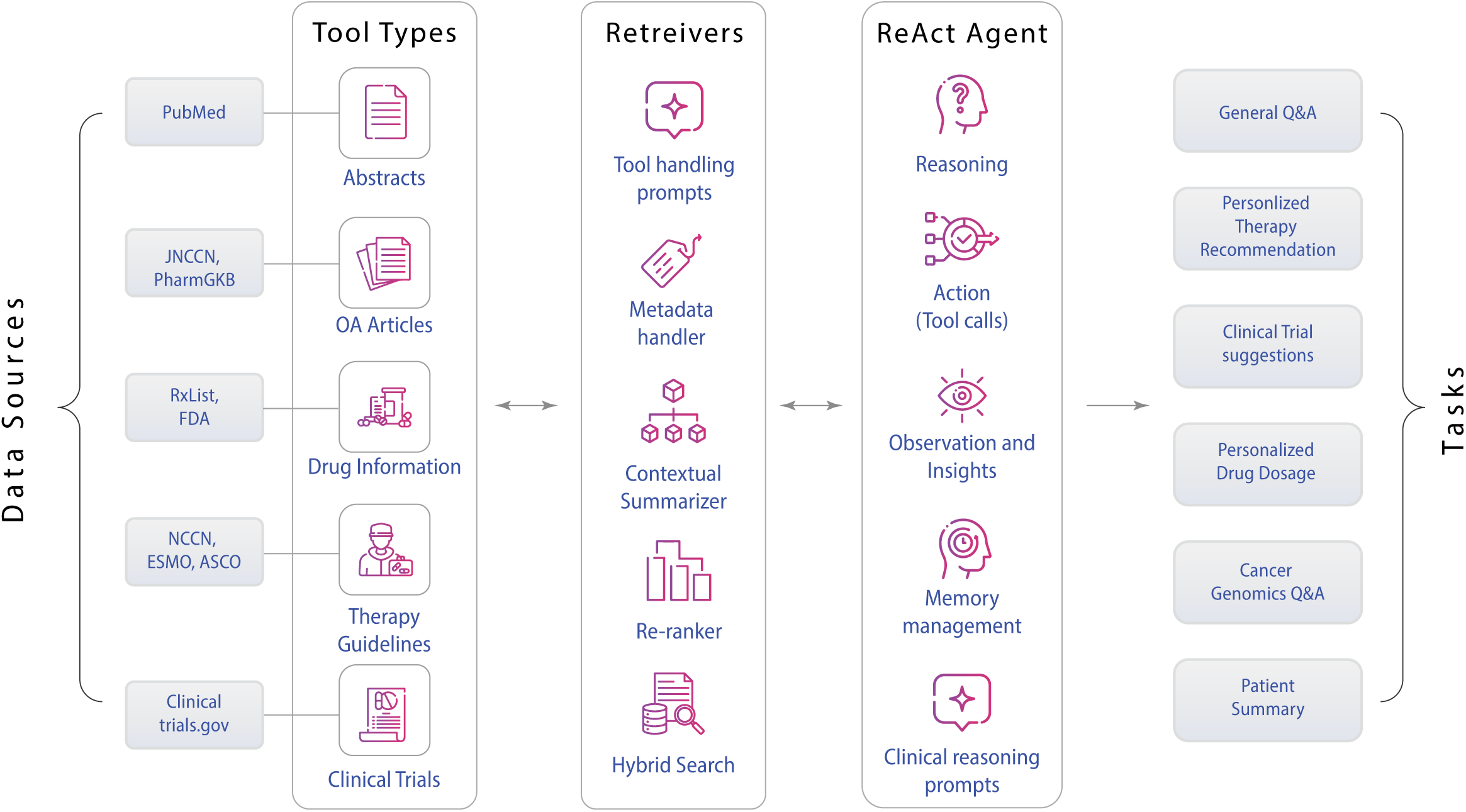
Illustration of the working of the GeneSilico CoPilot. Schematic workflow depicting the overall functionality of the agentic framework for precision oncology. The first step involves the collection and pre-processing of diverse medical data sources including literature, clinical trials, drug information and treatment guidelines – these serve as the tools. The second step involves the retrieval process where to efficiently extract relevant information given a query by employing appropriate tool selection, hybrid search, re-ranking, and summarization. Further, the information retrieved is fed into a ReAct Agent that implements a cycle of reasoning, action (tool calls), and observation to generate insights. The final step involves the synthesis of the response containing the medical insights and recommendations that caters to the use cases such as personalized therapy recommendations, clinical trial suggestions, genomic data analysis, and patient summaries.

For example, if tasked with recommending personalized therapies, the agent directs the appropriate tool to search the vector store. The tool retrieves and filters the results, selecting only the most relevant documents and generating insights, which are then passed to the agent for decision-making. Similarly, for drug dosage recommendations, a dedicated tool extracts and processes relevant data, providing refined insights for the agent to integrate into its reasoning loop (**Figure 2**).

**Figure 2:**
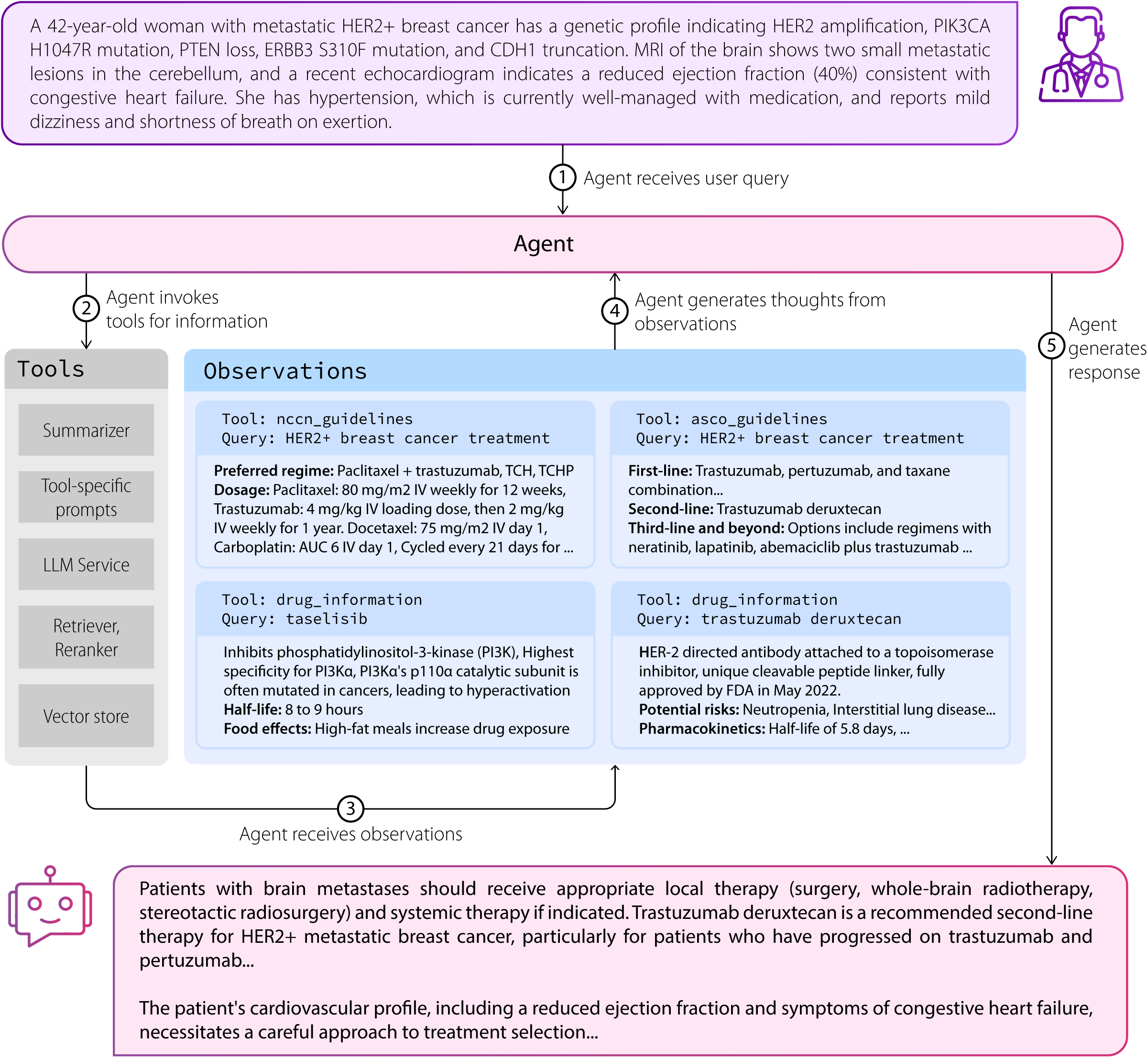
Flow of the process from a patient case to treatment plan. An oncologist provides a patient case study and prompts GSCP to recommend a treatment plan. Upon receiving this query, GSCP engages in a structured Reason-Action-Observation process and synthesize the response based on the patient’s specific clinical details. Step 2 through 4 runs in a loop till the agent has enough information to form a coherent response using its clinical reasoning abilities.

The GSCP employs a hybrid vector search strategy, combining dense embeddings (generated using Voyage AI) for capturing semantic relationships and sparse embeddings (using SPLADE) for extracting key features. The data corpus is categorized by source and use case, pre-processed, and siloed into collections, with each tool performing vector searches and tailored post-processing to ensure high-quality, relevant data retrieval.

### Systematic curation of relevant documents for the GSCP vector store

Vector databases rely on document embeddings for indexing and retrieval based on semantic similarity. However, semantically similar documents, even if thematically unrelated, can have close vector representations, leading to improper partitioning of the search space (**Figure 3a**). This often results in clustering documents from different sources together, particularly in specialized domains like breast cancer. Previously, naive chunking of long documents further complicated retrieval by fragmenting context, often breaking critical information connections across document segments.

**Figure 3:**
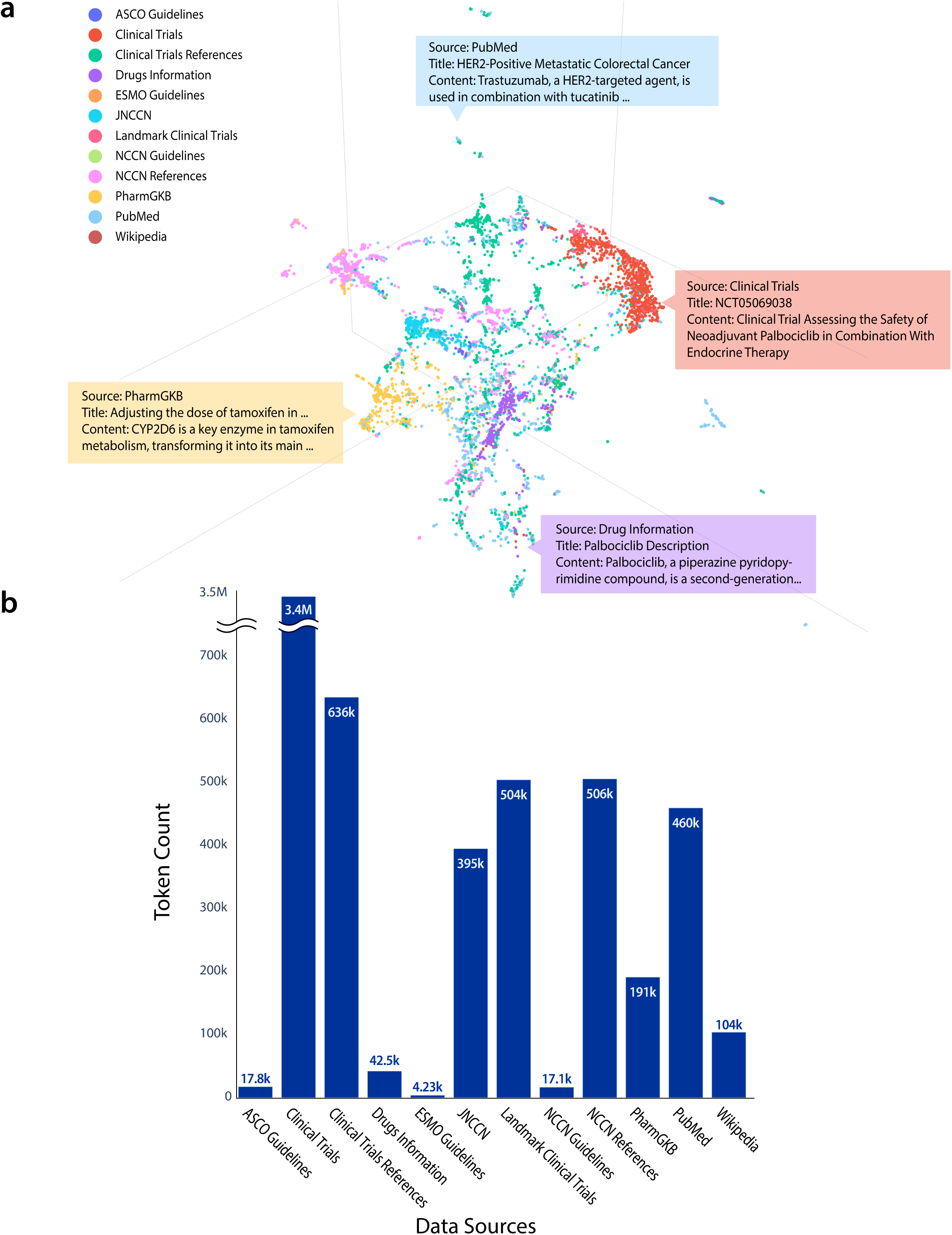
Representation of the data in the vector database. A) The U-Map in 3D to visualize the vector space that highlights the overlaps and intersections among various topics; B) The distribution of information across diverse topics within the vector database, illustrated through token counts, offers a comprehensive view of the content richness and topical breadth. Token count was generated using the `tiktoken` library.

In the proposed vector storage setup, we have eliminated chunking entirely, as modern LLMs can now accommodate entire documents in their context windows. This approach preserves the full document context, preventing the disruption caused by chunking and ensuring that essential relationships within the document remain intact. While this has addressed issues of context fragmentation, the embedding model is still limited by the number of tokens it can process. To resolve this, we now generate a summary of each document using LLMs and embed this summary. This ensures that the semantics are correctly encoded in the vector. During retrieval, both the full document and its corresponding summary, along with all the metadata are retrieved, allowing the agent to make use of both concise insights and the complete context of the document.

Our system also tackles the challenge of overlapping vector spaces that arise from thematically similar documents. For example, breast cancer guidelines from different sources (such as NCCN and ASCO) can have closely aligned content, leading to potential overlaps in the retrieval process. This issue is compounded by the imbalanced data volume across categories. For instance, breast cancer treatment guidelines from the National Comprehensive Cancer Network (NCCN) contain 17.1 thousand tokens, while clinical trial documents total 3.4 million tokens. The collated information from PubMed has a total of 460 thousand tokens. The volume discrepancies are illustrated in **Figure 3b**. This can bias retrieval, with smaller but crucial datasets being overshadowed by larger document sets. Despite being much smaller in volume, the guidelines and the generic information were used to synthesize a lot of responses during the evaluation process that is further illustrated in **Figure 4**.

**Figure 4:**
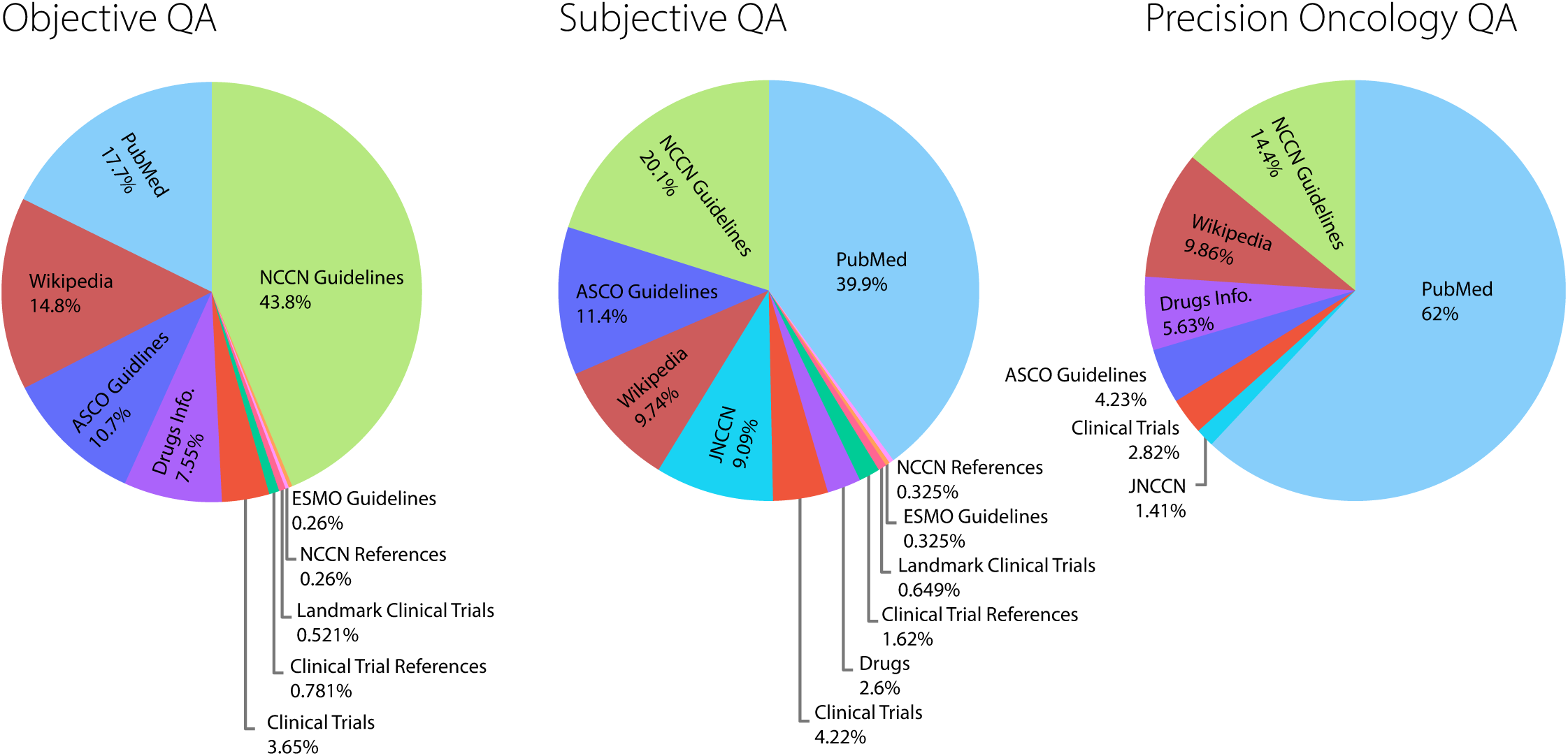
Representation of tool usage for diverse question-answering tasks. Tool usage distribution across three distinct question answering (QA) tasks: Objective QA, Subjective QA, and Precision Oncology QA (from left to right). In Objective QA, the NCCN Guidelines represent the largest reference source at 43.8%, followed by PubMed (17.7%) and Wikipedia (14.8%). Subjective QA shows a dominant reliance on PubMed (39.9%), with NCCN Guidelines (20.1%) and ASCO Guidelines (11.4%) also contributing significantly. In Precision Oncology QA, PubMed is the predominant source (62%), with NCCN Guidelines accounting for 14.1%. Each task demonstrates a distinct pattern of reference source utilization, with NCCN Guidelines and PubMed playing major roles across all categories.

To address these challenges, we propose a siloed abstractive vector storage system. This partitions the corpus into thematically distinct silos—such as NCCN Guidelines, ASCO Guidelines, PubMed, PharmGKB, Clinical Trials, and others. The storage of each silo is backed by a collection in the vector database. Each silo is managed by a dedicated retrieval module that applies tailored filtering, summarization, and processing techniques, ensuring that each source is retrieved and used independently. This prevents the intermixing of guidelines or documents from different sources, ensuring that responses are synthesized based on silo-specific information. For instance, responses that involve treatment guidelines are pulled exclusively from their relevant silo without contamination from other guideline sources.

With the elimination of document chunking, each silo document now includes only a summarized abstraction, rather than a chunked representation. These summaries are designed to be context-specific, based on the silo’s purpose. For example, a summary focusing on clinical trial eligibility criteria will differ from one that highlights key findings from a landmark drug trial, even though both originate from the same clinical trial document. This abstraction allows the agent to grasp document semantics more efficiently, reducing the number of reason-action loops required for response generation.

We utilize document summaries for both embedding and vector searches. Each document is summarized using an LLM, and the summary is encoded into a vector for efficient retrieval. During vector search, these encoded summaries are used to identify relevant documents. However, when retrieving, the system fetches the entire raw document, not just the summary, allowing the agent to have access to the full content.

After retrieval, the relevant portion of the full document is selected based on the query and used to generate insights and observations. These insights are crucial for the reasoning-action loop, where the agent synthesizes information to generate a comprehensive response. By leveraging the full document content and not limiting retrieval to just the summary, the agent ensures that all contextually significant information is considered. This approach maintains the coherence of the original document while also benefiting from the efficiency of summarized content during vector search. This strikes a balance between the computational efficiency of summary-based vector searches and the depth of information available from full-document retrieval.

### GSCP agent improves response quality over general purpose LLMs and RAGs

Agents act as a powerful approach for question answering tasks, particularly in the medical domain where access to comprehensive and informative answers is crucial. However, evaluating the effectiveness of these systems, especially in comparison to standalone LLMs or simpler RAG configurations, requires a rich and diverse set of evaluation datasets. Such datasets should encompass a variety of question formats, difficulty levels, and domains to provide a rigorous assessment of both context retrieval and response generation capabilities.

The absence of dedicated breast cancer question-answering datasets necessitated the creation of a comprehensive evaluation suite. We combined publicly available medical question-answering datasets with domain-specific samples encompassing both objective (multiple choice) questions from sources like MedMCQA^21^ and MedQA^22^, and subjective (open ended) questions from sources like PubMedQA^23^ and internally constructed questions, all related to breast cancer (**Figure 5**). For objective dataset, a Python script was utilized to identify all potential answer choices within the dataset for each question. Subsequently, we searched for the corresponding responses for the presence of these choices. This approach was essential as the correct answer was frequently not explicitly listed among the provided options, thereby simplifying the downstream evaluation task.

**Figure 5:**
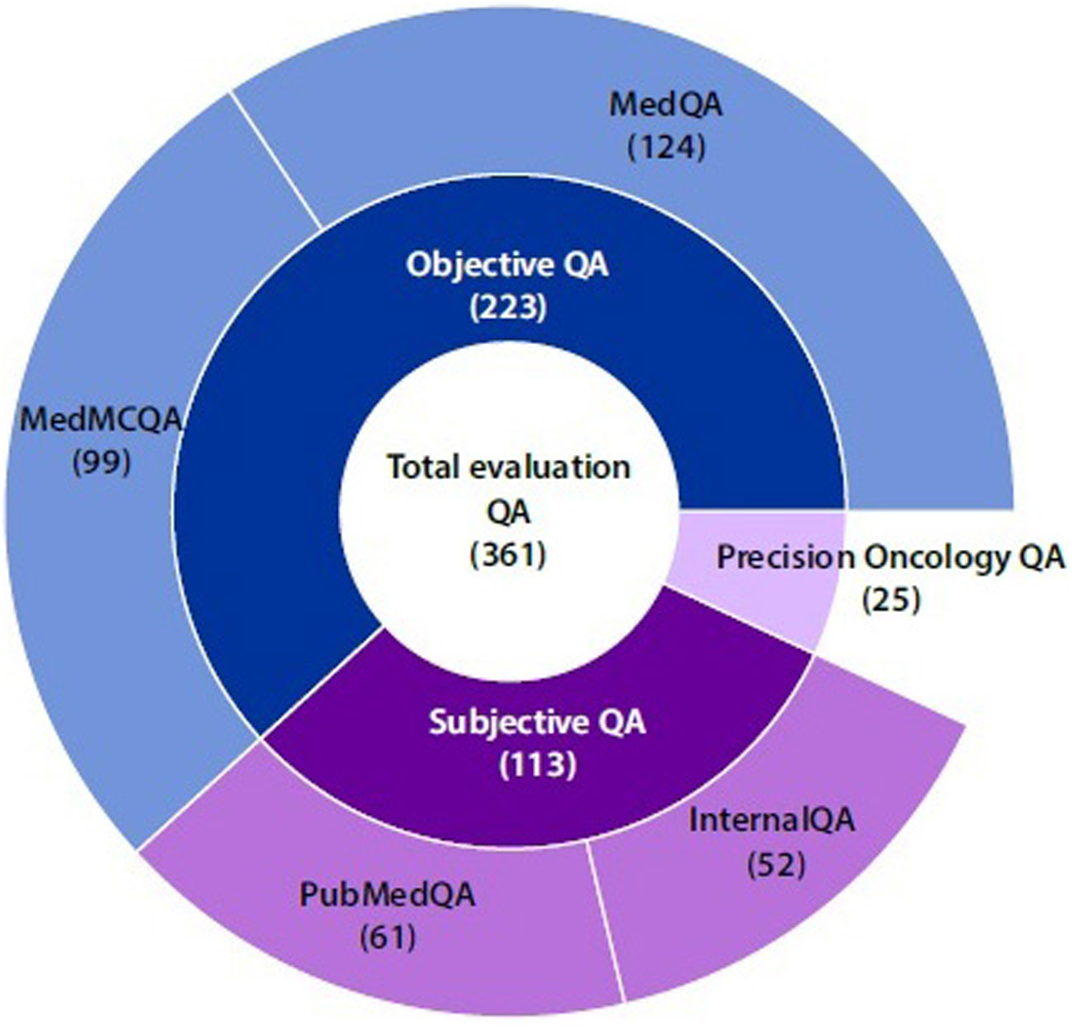
Distribution of the QA dataset. The donut chart presents the breakdown of 361 total questions across the different types of question answering tasks. Objective QA comprises 223 questions, sourced from *MedQA* (124 questions) and *MedMCQA* (99 questions). Subjective QA includes 113 questions, derived from *PubMedQA* (61 questions) and *InternalQA* (52 questions). Additionally, Precision Oncology QA, developed in-house dataset contributes 25 questions.

The application employs meticulously designed system prompts to enable the Reaction-Action or the ReAct loop as well as the thought and response generation process. The ReAct system prompt that is used here is a standard public-domain prompt that generates *“thoughts”*, *“actions”*, and *“observations”*. To capture the full complexity and subtleties of user query and generate detailed responses with evidence, we have an additional system prompt that is used in the intermediate stages of the response generation process. However, for evaluation purposes, this system custom prompt from the application is replaced to generate responses that aligns with the format of the evaluation to facilitate fair and consistent assessment. These prompts for evaluation are discussed in detail in the **Supplementary Information, Section 1**.

We employed state-of-the-art LLM services, GPT-4 and Claude Opus-3, known for their long context windows as well as their comparable performance on benchmark tests (**Figure 6**). The evaluation explored both simple RAG and agentic configurations.

**Figure 6:**
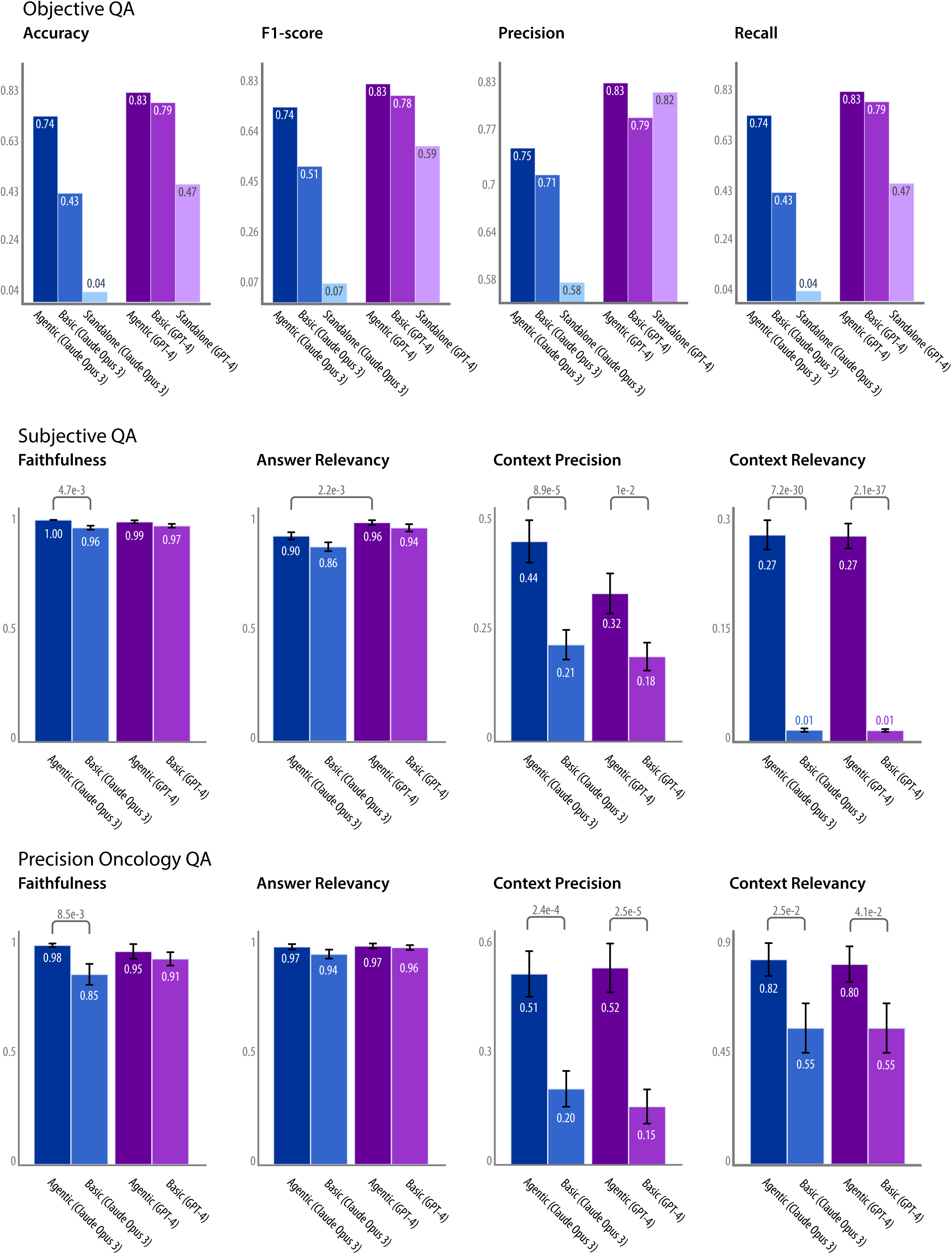
Assessment of the performance of GSCP on question answering tasks. Performance evaluation of GSCP across three different types of question answering (QA) tasks: Objective QA, Subjective QA, and Precision Oncology QA. The metrics used for Objective QA include Accuracy, F1-score, Precision, and Recall, while for Subjective QA and Precision Oncology QA, the performance is evaluated using Faithfulness, Answer Relevancy, Context Precision, and Context Relevancy. The bar plots provide a comparative view of these metrics across different datasets and models, highlighting the variation in performance.

For *Objective Question Answering* (QA) containing 223 questions, four quantitative metrics (accuracy, precision, recall, F1-score) were used to assess performance. Agentic systems significantly outperformed both RAG and standalone LLMs across all metrics. The GPT-4 powered agentic setup consistently achieved the highest scores in accuracy, recall, F1-score. Both LLM services demonstrated comparable performance, with GPT-4 exhibiting a slight edge. Agentic (GPT-4) consistently achieved the highest scores in accuracy, recall, F1-score, and precision (0.83, 0.83, 0.83, and 0.83 respectively), followed by the Agentic setup with Claude Opus 3 with moderate success. Basic RAG models showed mixed results, with relatively strong precision but lower accuracy, recall, and F1-score. Standalone LLMs exhibited the lowest performance across all metrics.

For *Subjective QA* consisting of 113 questions, the DeepEval framework evaluated retrieval and generation performance for subjective questions. Standalone LLMs were excluded due to the absence of a retrieval context in their responses. Context precision, context relevancy, faithfulness, and answer relevancy metrics were employed. Agentic systems achieved superior performance in both retrieval and generation tasks. In terms of context precision, *Agentic (Claude Opus 3)* led with a score of 0.44, followed by *Agentic (GPT-4)* at 0.37, while basic models struggled. Both Agentic models achieved high context relevancy scores of 0.27, significantly surpassing the basic RAG models. For answer relevancy, *Agentic (GPT-4)* excelled with a score of 0.96, followed by *Agentic (Claude Opus 3)* at 0.90, while basic RAG models performed reasonably well. Finally, while faithfulness scores were comparable overall, *Agentic (Claude Opus 3)* achieved a perfect score of 1.

A custom in-house dataset focusing on precision oncology and breast cancer genetics, containing 25 questions, was created to simulate real-world healthcare complexities. This dataset, *Precision Oncology QA*, mimicked genetic markers, disease progression, and personalized treatment options. The same metrics used for subjective questions were applied. Agentic systems significantly outperformed basic systems in both context precision and relevancy. *Agentic (Claude Opus 3)* achieved a precision score of 0.51 and a relevancy score of 0.82, while *Agentic (GPT-4)* scored 0.52 for precision and 0.80 for relevancy. In contrast, basic RAG systems showed lower scores, with *Basic (Claude Opus 3)* at 0.20 for precision and 0.55 for relevancy, and *Basic (GPT-4)* at 0.15 for precision and 0.55 for relevancy. While answer relevancy and faithfulness scores were comparable across models, *Agentic (Claude Opus 3)* demonstrated slightly higher faithfulness with a score of 0.98 compared to *Basic (Claude Opus 3)* at 0.85. These evaluations demonstrate that moving from a RAG configuration to the proposed Agentic setup will improve performance no matter which LLM service is being used.

To construct a faithful representation of the clinical setting, synthetic patient case studies were generated through a collaborative effort involving practicing oncologists and LLM services. Oncologists contributed essential clinical insights, ensuring the case studies accurately reflected real-world medical complexities. These expert-provided details, devoid of specific patient data, served as the foundation for the LLMs to craft comprehensive case studies. To further enhance the authenticity of these synthetic cases, oncologists were asked to review the generated case studies to validate their alignment with the real clinical reports. The performance of agentic models, specifically Claude Opus 3 and GPT-4, was assessed using these four synthesized patient health records. In these experiments, the models were tasked with formulating suitable treatment plans based exclusively on the presented patient data. The detailed model outputs along with can be found in **Supplementary Information Section 2**.

### Decoding the coordination among tools that improve GSCP response quality

A key strength of the GeneSilico Copilot (GSCP) agent lies in its ability to intelligently coordinate a suite of specialized tools for retrieval, processing, and reasoning across distinct topics. Each tool is responsible for accessing and processing content from specific silos, such as NCCN guidelines, PubMed, and clinical trial data, based on the nature of the query. Every tool along with their descriptions and number of documents that are retrieved from that silo is present in **Supplementary Information, Section 3**. This segmented approach ensures that the tool retrieves the most relevant information for a given query while maintaining transparency in the agent’s decision-making process. By exposing its reasoning steps, the agent provides insight into how different sources contribute to the final response, allowing the end-user to better understand the weight and relevance of each information source (**Supplementary Figure 1**).

**Figure 7** breaks down the tool utilization over reasoning steps across different evaluation scenarios. For instance, in objective case study questions, the agent heavily relies on NCCN guidelines, which are critical for treatment protocols and recommendations. Meanwhile, PubMed is consistently utilized as a supplemental tool to enrich responses with broader biomedical literature. This balance between targeted guidelines and general literature highlights the flexibility of the agent in adapting to various types of questions and datasets. The coordination process typically occurs within a sequence of 3 to 5 reasoning steps, as shown in the **Figure 7**. In each step, the agent identifies the relevant tool while the tool retrieves information, integrates the insights, and helps the agent in evolving the response. This breakdown not only demonstrates the logical flow of the agent’s reasoning but also showcases its ability to adaptively select the most appropriate tool based on the specific needs of the query. The tool orchestration within this siloed architecture significantly enhances the quality and precision of the responses generated, providing a powerful mechanism for clinical decision support (**Supplementary Figure 2**).

**Figure 7:**
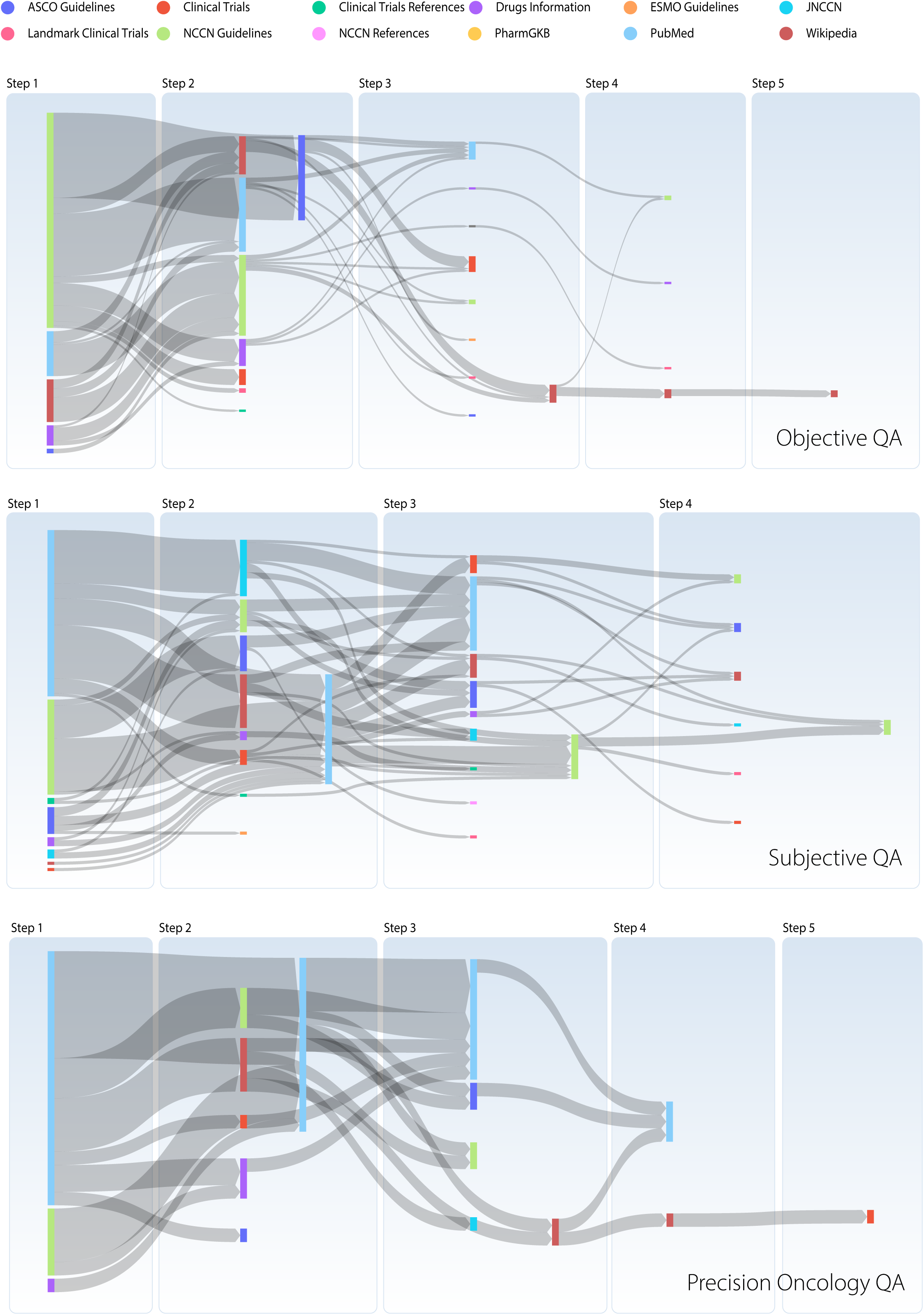
Methodical use of tools in step-by-step response synthesis. The Sankey diagrams illustrate the utilization of various tools across multiple steps in the response generation process for Objective QA, Subjective QA, and Precision Oncology QA tasks. Each chart represents the flow of tool usage from Step 1 through to Step 5, detailing the number of steps taken to generate responses and the specific tool used at each stage.

## DISCUSSION

Large language models have demonstrated considerable potential across various domains, including healthcare and biomedical research. However, limitations in transparency and robust evaluation methodologies have hindered their full clinical integration. GeneSilico CoPilot (GSCP) addresses these challenges by proposing an agent-based framework that leverages the inherent reasoning capabilities of LLMs to plan and execute tasks within the healthcare domain. This work focuses on the specific domain of breast cancer, showcasing the advantages of the GSCP framework over standalone LLMs and Retrieval-Augmented Generation (RAG) systems in both generic question answering and precision oncology tasks.

A key differentiator of GSCP, when compared to commercially available language models is its ability to expose the reasoning process. Once the user provides a query, the agent, as instructed by system prompts, creates clinically relevant thoughts, actions and observations. Every observation is created with in-text citations. Users can directly go to the URLs to verify the content of the observations from the tools. Furthermore, while not all parts of the observations may be necessary in forming the final response, the observations themselves act as crucial supplementary information. The final response also provides URL references, improving the trustworthiness of the system.

Evaluations conducted across public and private datasets demonstrate the superiority of the proposed agent-based framework compared to traditional RAG systems. In precision oncology question answering, GSCP achieved an improvement of up to 15.29% in answer faithfulness compared to RAGs. Retrieval metrics also showed significant improvement, with GSCP system achieving up to 200.83% and 47.27% better performance in context precision and context relevancy, respectively. These results highlight the clear advantage of the agent’s reasoning and retrieval mechanisms over basic RAG approaches. Similar improvements were observed in the subjective question answering dataset, where the GSCP agent achieved up to 93.65% and 2600% improvement in context precision and context relevancy, respectively. The agent’s retrieval mechanism facilitates a more robust reasoning process, and by incorporating these reasoning steps into the response generation, GSCP system enhances the trustworthiness and transparency of its answers.

The GSCP framework surpasses the limitations of simple RAG systems by employing a highly specialized bespoke ReAct agent that integrates breast cancer-specific clinical rationale into the reasoning loop. The agent leverages a dynamic prompt selection mechanism to enhance its ability to process and synthesize information across various silos of biomedical data.

Furthermore, GSCP employs a suite of specialized tools, each designed to interact with specific types of data silos—such as guidelines, clinical trials, and biomedical literature. Each tool call is triggered by the ReAct prompt but powered by the custom retrieval prompt that changes based on the user query and the silo, which ensures that the tool extracts relevant information from the documents. The tool uses LLMs to assess the retrieved documents based on the agent’s query, generating observations that contain evidence, citations, and insights. The insights are critical, as they provide statements that logically support the user query, contributing to the reasoning flow of the agent. The agent’s final response is an outcome of the logical synthesis of these observations. It uses the custom system prompt to generate a comprehensive and coherent response, incorporating the relevant insights and evidence. This step ensures that GSCP agent produces detailed, evidence-backed responses, offering clinicians greater confidence in the accuracy and relevance of the system’s output. This approach represents a significant evolution over traditional RAG systems, improving both the depth and precision of the agent’s responses.

Our evaluation revealed that Claude Opus 3 produced well-structured responses that resonated with oncology experts, despite achieving lower overall evaluation scores compared to OpenA’s GPT-4. While Opus 3 exhibited slower response generation times, often requiring up to three minutes to complete a response using provided tools, its outputs were characterized by superior readability. In terms of medical accuracy, Opus 3 offered more detailed explanations, including comprehensive drug and dosage information, while GPT-4 produced simpler responses.

Although both systems demonstrated comparable levels of medical accuracy, the significant disparity in human-perceived readability suggests an inability of the DeepEval evaluation framework to fully capture the nuanced aspects of response quality, particularly when considering human factors such as readability. This finding underscores the limitations of relying solely on automated metrics to assess model performance, particularly in complex domains such as medicine. While medical accuracy is undeniably crucial, it is essential to recognize that it is not the sole determinant of response quality. A comprehensive evaluation should consider additional factors, such as response clarity, coherence, and overall clinical utility, as perceived by human experts.

The GSCP system’s transparent planning process, which can be visualized through tool usage, provides valuable insights into the relative importance of information sources. For example, our observations indicate a clear preference for NCCN guidelines over American Society of Clinical Oncology (ASCO) and European Society for Medical Oncology (ESMO) guidelines. It is noteworthy that NCCN guidelines underwent a meticulous manual paraphrasing process to convert them into plain text while preserving the information conveyed in the original flowcharts. In contrast, ASCO and ESMO guidelines primarily relied on LLM-based summarization. PubMed also emerged as a significant information source. While PubMed offers a wealth of open-access articles containing general knowledge, our focused initial retrieval process effectively transformed the PubMed collection into a more specialized corpus tailored to the domain of oncology. Analysis of tool usage statistics can be leveraged to inform future optimizations of the data sources, potentially leading to the deprecation, consolidation, or replacement of certain sources based on their effectiveness within the agent’s framework.

Future endeavours include expanding our testing to encompass real-life patient cases and evaluating the GSCP system’s capabilities in therapeutic decision support. This necessitates the development and implementation of robust and reproducible evaluation metrics. Current frameworks like DeepEval, which rely on LLM services for evaluation, are susceptible to inconsistencies. Therefore, there is a pressing need for more sophisticated evaluation methods with human-in-the-loop specifically designed to assess the planning and reasoning capabilities of LLMs.

The GSCP system currently faces some limitations in terms of processing speed. The tool usage and frequent communication with the LLM service contribute to a processing delay, with complex cases requiring up to five minutes for final response generation.

Developing a patient-specific treatment regimen requires meticulous evaluation of various factors, including the patient’s medical history, comorbidities, prior treatments, and potential drug toxicities. This necessitates a comprehensive review of the patient’s medical records, encompassing laboratory results, imaging studies, and medication history. A thorough understanding of the patient’s current health status and any coexisting conditions is also essential. Once this data is collected, the physician can begin exploring treatment options aligned with established clinical guidelines from NCCN, ASCO, and ESMO. These guidelines provide evidence-based recommendations informed by the latest research and clinical experience. However, it is equally important to consider the patient’s individual needs and preferences, as well as their eligibility for ongoing clinical trials offering access to potentially groundbreaking therapies. This complex decision-making process necessitates the synthesis of information from diverse sources. The GSCP system addresses this challenge by leveraging its knowledge base to recommend personalized treatment plans for each patient case. This streamlines the physician’s workflow, facilitates informed decision-making, and ultimately contributes to enhanced patient care.

In conclusion, this work demonstrates the potential of developing domain-specific agentic systems. By focusing on a particular domain, such as oncology, the system can be optimized to effectively process and generate information within the context of a vast and complex data landscape.

## METHODS

### Data sources

GSCP leverages a collection of manually curated data sources specific to breast cancer, compiled with the support of practicing oncologists. These sources include standard breast cancer guidelines from the National Comprehensive Cancer Network (NCCN), American Society of Clinical Oncology (ASCO), and European Society for Medical Oncology (ESMO).

#### Targeted Drug and Gene Information

To incorporate relevant drug and gene information, a curated list of 68 genes (including HRR and pharmacogenomics genes) and their targeted drugs was compiled. A customized GeneSilico gene panel for breast cancer therapy recommendations was designed, encompassing these 68 genes. The selection criteria for these genes included: genes associated with therapies (FDA-approved, Phase 3, and Phase 4 clinical trials); genes with high research significance and frequent alterations in databases like Human Somatic Mutation Database (HSMD) (digitalinsights.qiagen.com/hsmd/) and cBioPortal (www.cbioportal.org); genes associated with homologous recombination repair (HRR) mechanism; pharmacogenomic (PGx) genes relevant to breast cancer; normalized codon length of genes; and key genes present in other somatic panels such as MSK-IMPACT (www.mskcc.org/msk-impact), Foundation Medicine CDx diagnostic panel (https://www.foundationmedicine.in/our-services/cdx.html), and MedGenome panel (diagnostics.medgenome.com). The rankings from these criteria were combined using a rank aggregation algorithm to determine the final list of top genes, which were then manually validated. Pathogenic and likely pathogenic variants in breast cancer were selected using the HSMD, COSMIC^24^, and ClinVar^25^ databases. Additionally, the GeneSilico gene panel for breast cancer includes all exonic and a few selected intronic variants for the 68 genes as well as 32 microsatellite instability (MSI) hotspots.

Subsequently, this targeted drug-gene list was used to extract drug data from Drugbank Open Data (go.drugbank.com/releases/latest#open-data) ^26^, FDA drug labels (labels.fda.gov), RxList (www.rxlist.com), Therapeutic Target Database^27^, Drugs.com (www.drugs.com), and Wikipedia. Drug approval details were obtained from the FDA and ClinicalTrials.gov. We used the OpenFDA API and the Clinical Trials API to access the information. PubMed information was gathered using the PMC E-Utilities. PharmGKB and JNCCN provided further breast cancer-specific literature.

#### Data Abstraction and Summarization with Contextual Focus

To enhance context for lengthy documents, all documents were summarized using LLM services. Instead of generic summaries, task-specific summaries were created. This context-aware process facilitated the summarization of pertinent sections rather than entire documents. The summarization was performed using Google Gemini 1.0 Pro. The summarization prompt was provided depending on the requirement. For summarization of clinical trials for eligibility criteria, the prompt was `“Make the following clinical trial information concise, highlighting the key eligibility criteria. Simply respond with the shortened text in markdown format.”` For clinical trials which contained drug approval information, the prompt was changed to `“Give an abstract of the trial highlighting the drug approval information. Simply respond with the shortened text in markdown format.”` In every case, the summary was formatted with markdown tags. In our experiments, using markdown tags improved the retrieval quality. This allowed the tools to fetch fewer documents and helped optimize the context window use. Although storing documents with multiple summaries creates redundancy across silos, this approach enhanced agent performance. This summarization process was applied to all data sources containing long-form textual content.

#### Data Staging for Manageability and Retrieval

Data staging was implemented to improve manageability and retrieval efficiency. Before embedding, a copy of the data, along with extracted metadata, summaries, and URLs (for public domain documents), was stored in a NoSQL datastore. This was implemented using MongoDB. This approach simplifies and automates the embedding process. The stored URLs and metadata are utilized by system prompts for response refinement.

#### Manual Curation for Complex Documents

Documents such as NCCN guidelines containing complex diagrams and flowcharts, underwent manual paraphrasing and conversion into plain text while preserving the step-by-step narrative. Furthermore, NCCN documents were segmented based on cancer subtype, treatment phase, and treatment nature. Most other documents, including guidelines from ASCO and ESMO, were summarized using LLMs followed by manual inspection. Each summary was further segmented and annotated with markdown tags to enhance the agent’s contextual understanding and facilitate the generation of more relevant responses.

### Vector stores

In our vector store implementation, we employ a hybrid search strategy that leverages both dense and sparse vector embeddings to enhance retrieval performance. Specifically, we generate and store two types of embeddings for each document: a dense embedding provided by a closed-source algorithm from VoyageAI, and a sparse embedding generated using SPLADE. During retrieval, we utilize a multi-stage process involving sparse vector matching, dense vector search, and Reciprocal Rank Fusion (RRF) to produce the final ranked list of documents.

#### Embedding Generation

For the dense embeddings, we use a proprietary model from VoyageAI that maps each document summary ***d*** and query ***q*** into a 1536-dimensional dense vector space. The embeddings are represented as:

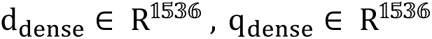

These embeddings capture the semantic representations of the documents and queries in a continuous vector space, facilitating similarity computations based on their geometric properties. Even though the document summaries are encoded, the summaries themselves contain the semantics of the whole document and therefore, should contain the semantics of the entire document.

Similarly, the sparse embeddings are generated using SPLADE, which produces high-dimensional sparse vectors reflecting the term importance within the vocabulary space. Let ***N*** denote the size of the vocabulary. The sparse embeddings for documents and queries are:

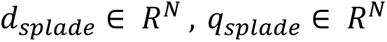

Here, N = 30522. Each element *d_splade,i_* or q_splade,i_ corresponds to the importance of the *i^th^* term in the vocabulary for the document or query, respectively. The sparsity of these vectors ensures computational efficiency during matching.

#### Storage of embeddings

Both the dense and sparse embeddings are stored using 16-bit floating-point precision (float16) to maintain a balance between storage efficiency and computational accuracy. This dual-storage approach allows us to utilize the strengths of both embedding types during retrieval. To facilitate efficient similarity searches during retrieval, we index the embeddings using the Hierarchical Navigable Small World (HNSW) algorithm, implemented via Qdrant. The HNSW index is particularly well-suited for high-dimensional data, making it an ideal choice for our dense embeddings of dimension 1536 and sparse embeddings corresponding to the vocabulary size. The HNSW index is configured with the following parameters:

m=16: This parameter determines the number of bi-directional links created for each element during the construction of the HNSW graph. A higher m value increases the connectivity of the graph, potentially improving search accuracy but also increasing memory consumption and indexing time. By setting m=16, we achieve a balance between index performance and resource utilization.

ef_construct=100: This parameter controls the size of the dynamic list of nearest neighbors during the index construction phase. A larger ef_construct value leads to a more accurate and robust index at the cost of longer indexing time. With ef_construct=100, we enhance the quality of the index without incurring excessive computational overhead.

full_scan_threshold=10000: This threshold defines the dataset size below which the system will perform a full brute-force scan instead of using the HNSW index. For datasets smaller than 10,000 vectors, a full scan is often more efficient due to the overhead associated with indexing. This ensures that we optimize retrieval performance across different dataset sizes.

#### Retrieval

The retrieval process consists of three main stages: sparse vector pre-fetching, dense vector searching, and Reciprocal Rank Fusion. Initially, we pre-fetch ***n*** candidate documents from the corpus using sparse vector matching based on the SPLADE embeddings. The matching score between a query ***q*** and a document ***d*** is computed using an inverse document frequency (IDF)-weighted inner product:

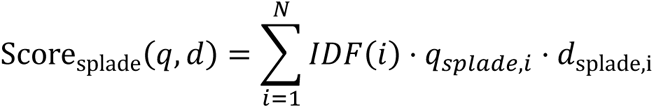

Here, *IDF(i)* is the inverse document frequency of the i-th term, which emphasizes the significance of less frequent terms in the matching process. The top n documents with the highest Score_splade_(*d*, *q*) are selected for further processing. Within the pre-fetched set of *n* documents, we perform a dense vector search to retrieve the top *m* documents m < n that are most semantically like the query. The similarity between the dense embeddings of the query and a document is calculated using cosine similarity:

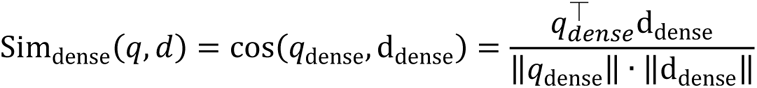

The documents are ranked based on Sim_dense_(*d*, *q*) and the top m documents are selected for the final stage. To combine the strengths of both the sparse and dense retrieval methods, we apply Reciprocal Rank Fusion (RRF) on the m documents obtained from the previous stage. RRF is an effective method for aggregating rankings from different sources by assigning higher scores to documents that appear near the top of multiple rankings.

Let *r_splade_*(*q*) be the rank of document *d* in the initial sparse ranking and *r_splade_*(*q*) be its rank in the dense ranking. The RRF score for each document is computed as:

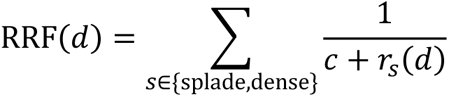

where c is a constant to control the impact of the rank positions. The documents are then sorted based on their RRF scores, and the top *k* documents *(k<m)* are selected as the final retrieval results. The values of *n=10*, *m=7*, and *k=5* is chosen based on empirical performance evaluations, ensuring that *n>m>k* to progressively refine the candidate set.

### The ReAct agent

The GSCP employs a bespoke ReAct agent, designed to simulate clinical reasoning processes for precision oncology decision-making. The agent is built ground up using the LlamaIndex’s *Workflows*, an event-driven architecture. Based on the general ReAct architecture but has been significantly modified to address the unique requirements of clinical workflows.

#### ReAct Architecture Overview

The ReAct architecture is a framework that enables agents to perform complex reasoning and action sequences by iteratively generating thoughts and actions based on user inputs. In this architecture, a system prompt guides the agent to utilize a set of tools to achieve the desired outcome. The workflow begins with the agent receiving a user query. The agent then formulates a thought on how to address the query, which often involves selecting an appropriate tool to use. The action is the execution of this tool, and the result is an observation that informs the agent’s next thought. This thought-action-observation loop continues until the agent arrives at a final response, effectively solving the user’s query through a combination of reasoning and tool usage. The initial system prompt, referred to as the ReAct system prompt, provides general instructions to the agent on how to perform these steps.

#### Modifications for Clinical Reasoning

While the standard ReAct prompt is effective for general-purpose reasoning, it lacks the specificity required for clinical reasoning in oncology. To address this, we introduced a second system prompt, the *clinical reasoning system prompt*. This specialized prompt takes precedence over the default ReAct prompt during key stages of the agent’s workflow, particularly those involved with thought formation and output generation from observations.

This override is essential during these stages because clinical decision-making, especially in complex fields like oncology, demands more than basic reasoning. The agent must process detailed patient data, cross-reference clinical guidelines, and synthesize information from diverse sources such as clinical trials and drug information databases. By using a specialized prompt, the agent can:

- Simulate the collective clinical reasoning typically exhibited by a virtual tumour board, considering factors like patient history, disease progression, and personalized treatment options.
- Ensure that the thoughts generated by the agent are aligned with clinically appropriate logic, driving the selection of the most relevant tools and observations.
- Prioritize and refine observations based on the established clinical standards, making the agent’s decision-making loop more suited to precision oncology tasks.

#### Agent Workflow

The ReAct agent’s workflow is broken down into four distinct functions:

1. Get User Query: The agent receives the user’s input, typically a clinical question or patient case, which forms the starting point of the reasoning process. This is where the ReAct system prompt is also introduced.
2. Prepare the Chat History: The agent structures the previous interactions and context into a coherent history, ensuring it can build upon prior information effectively. This is where the *clinical reasoning system prompt* takes over.
3. Generate Tool Calls (Create Thought): Now, using the *clinical reasoning system prompt*, the agent forms a thought and decides which tool to call. This thought is shaped by clinical logic, guiding the selection of appropriate tools for the query.
4. Process Tool Calls (Get Observations): The selected tool returns observations, which are integrated back into the reasoning loop. This also collates and generates the citations.

Steps 2, 3, and 4 are repeated iteratively until the final thought leads to a comprehensive response.

#### Iterative Reasoning with Tool Feedback

Each tool used by the agent operates with its own *tool system prompt*, which follows a shared template but is customized based on the tool’s function. The tools include modules for retrieving clinical trial data, drug information, precision oncology guidelines, and general literature searches. The tool system prompt is tasked with generating a summary relevant to the agent’s current query, given the documents or data it has accessed. This process involves distilling complex medical information into actionable insights that the agent can use in its reasoning loop. Depending on the tool’s nature, the prompt may instruct the tool to perform a tree-summarization—breaking down information into hierarchical components—or to compact or refine information to highlight the most critical elements. As the agent processes observations from the tools, it continually refines its thoughts and actions. The *clinical reasoning system prompt* guides this iterative process, encouraging the agent to consider alternative hypotheses, weigh evidence, and explore different facets of the patient’s case. This iterative loop continues until the agent synthesizes a comprehensive response that reflects a deep understanding of the clinical scenario.

#### Incorporation of Citations and Metadata

An essential aspect of clinical reasoning is the ability to reference authoritative sources. Both the ReAct and clinical reasoning system prompts instruct the agent to generate citations from metadata associated with the retrieved documents. Our vector store is constructed to include URLs and other reference information, allowing the agent to provide precise citations at each step of its reasoning. This feature not only adds credibility to the agent’s responses but also enables users to verify and further explore the referenced material.

#### Memory Architecture

To handle complex medical queries while efficiently managing the limited context window, the ReAct agent uses two distinct types of memory:

1. **Conversational Memory**: This memory stores the user’s messages and the final agent responses. It handles the primary conversation between the agent and the user, ensuring that important interactions are preserved for continuity without overloading the context window.
2. **Working Memory**: This memory tracks the internal conversations between the agent and the tools. It stores the agent’s tool-specific queries and the corresponding observations, keeping these interactions separate from the user conversation. By isolating the tool interactions, the agent can manage detailed queries and observations while freeing up space in the context window for ongoing user interactions.

This separation of memory ensures efficient management of complex interactions, but it comes with one limitation: the user cannot directly reference or ask questions about tool-generated observations since these observations are stored in working memory. This drawback can prevent the user from delving into specific details generated by a tool unless the agent explicitly includes them in the final response. This highlights a limitation in flexibility when it comes to user-driven exploration of tool-generated insights.

#### Context Overflow Handling

In scenarios where the response generation requires numerous reasoning loops and observations begin to quickly fill up the context window, GSCP employs insight extraction and compression to manage context overflow efficiently specifically within the working memory. This process ensures that the agent can continue its reasoning tasks without losing critical information or running out of context window capacity. Here, we compress the middle portion of the conversation while preserving the most critical parts—the initial user input and the current reasoning step. When the content of the current working memory has exceeded the context window, we then use another instance of the same LLM to create a compressed representation of the past observations by extracting the necessary insights. By compressing the middle sections of working memory and retaining the most important insights and final observations, the agent ensures that it can continue its reasoning process without disruption. The final response is generated based on these insights, ensuring a comprehensive and logical outcome.

### Experimental setup

We evaluated the proposed method using datasets constructed from public sources and real-life cases. Standard public datasets for breast cancer are unavailable. Therefore, we extracted breast cancer-related questions from multiple sources and categorized them as subjective (requiring long-form answers) or objective (multiple-choice). A simple keyword-based search facilitated extraction, followed by manual review by practicing oncologists to ensure question correctness. The objective dataset comprised 223 questions from MedMCQA and MedQA (USMLE), while the subjective dataset consisted of 113 questions extracted from PubMedQA and an in-house dataset (InternalQA). For objective questions lacking a single clear answer where oncologists identified multiple correct options, the questions were reworked as subjective ones. To simulate the complexities encountered by medical professionals in real-world oncology practice, we constructed a custom in-house dataset, the Precision Oncology dataset, of 25 questions focused on precision oncology and breast cancer genetics. This dataset embodies case-study like scenarios, mimicking an oncologist’s investigative process. The internal dataset as well as the precision oncology datasets were created with the support of practicing oncologists. The dataset for evaluation is provided in **Supplementary Data**.

Accuracy, F1 score, precision, and recall were used to assess system performance for simple multiple-choice questions. For the subjective and precision oncology datasets, the DeepEval framework (https://docs.confident-ai.com/) evaluated our system and compared its performance to a RAG system. This framework employs Contextual Precision (ranking relevant information), Contextual Relevancy (overall retrieved context relevance), Faithfulness (factual alignment between response and retrieved context), and Answer Relevancy (ratio of relevant statements in the answer) to measure the retrieval and generation performance.

## Data Availability

This is already mentioned in the Data Availability section of the manuscript.

## DATA AVAILABILITY

The datasets used in this study are available in the supplementary materials. **Supplementary Data** file contains evaluation datasets, including the objective questions extracted from MedMCQA and MedQA (USMLE), subjective questions from PubMedQA and our in-house dataset (InternalQA), and custom Precision Oncology dataset. Our in-house datasets (InternalQA and Precision Oncology) were created with the support of practicing oncologists and are included in the supplementary materials. The DeepEval framework used for performance evaluation is publicly accessible at https://docs.confident-ai.com/. Any additional data that supports the findings of this study are available from the corresponding author upon reasonable request.

## SUPPLEMENTARY INFORMATION

### Files

#### Supplementary Information

**Section 1:** Contains description of the datasets present in Supplementary Information Data and the prompts used to generate the responses for the evaluation.

**Section 2:** Contains selected synthetic case studies for intervention plan. These case studies were designed by oncologists based on real-life cases. The file contains the entire response provided by GSCP when presented with the case studies.

**Section 3:** Contains the configuration file of the different tools used by the agent. The “top k” and the “sparse k” determine the number of results fetched based on the dense and the sparse vector match respectively.

**Supplementary Figure 1:** Implementation of the agent.

**Supplementary Figure 2:** A visual explainer of the reasoning process for a case study.

#### Supplementary Data

Excel spreadsheet consisting of the questions and the corresponding ground truth for the datasets – Subjective, Objective and Precision Oncology.

## Competing interests

A provisional patent has been filed (Attorney Docket No. 107881-00005) describing the agentic framework. All the authors, except for SB are GeneSilico. Inc. employees/partners.

## ACKNOWLEDGEMENTS

The authors would like to thank Claire Cohen from GeneSilico Inc., and Ruhani Bhatia for their valuable input.

## AUTHOR CONTRIBUTION STATEMENT

DS conceived the study. RD and KM designed and implemented the agent and performed all experiments under the supervision of DS. SS, JN, and SM defined the scope for gene panel, and data ingestion. JN and SS also performed evaluations and internal checkpoints for review. JN generated diagrams. AP also implemented DeepEval. NA implemented the vector database and MongoDB, contributed to the agent design, and led the productionization efforts. AP assisted with vector database implementation, conducted data scraping and collation, and developed tools for agent use. VU and AS developed and deployed the copilot web application in production, with support from TS and HC. RT, SP, and SN provided clinical input to ensure the relevance of data and outputs. RT further contributed to the design of clinical questions and responses. BG, UM, SB, and MF provided critical feedback on the agent design. All authors reviewed and approved the final manuscript.

## Notes

### Competing Interest Statement

A provisional patent has been filed on 10/02/2024 (DW No.: 107881-00009, Serial No.: 63/702,408) describing the agentic framework. All the authors, except for SB are GeneSilico, Inc. employees/partners.

### Funding Statement

The entire study is funded by GeneSilico, Inc.

### Author Declarations

The study used openly available human data sources such as Pubmed, PharmGKB, Clinical Trial databases, DrugBank Open Data, and NCCN/ASCO/ESMO.

### Summary of Updates

Contains updated methodology section with more details. Supplementary section is also updated.

