## Supplementary Figures for "Improved precision oncology question-answering using agentic LLM"

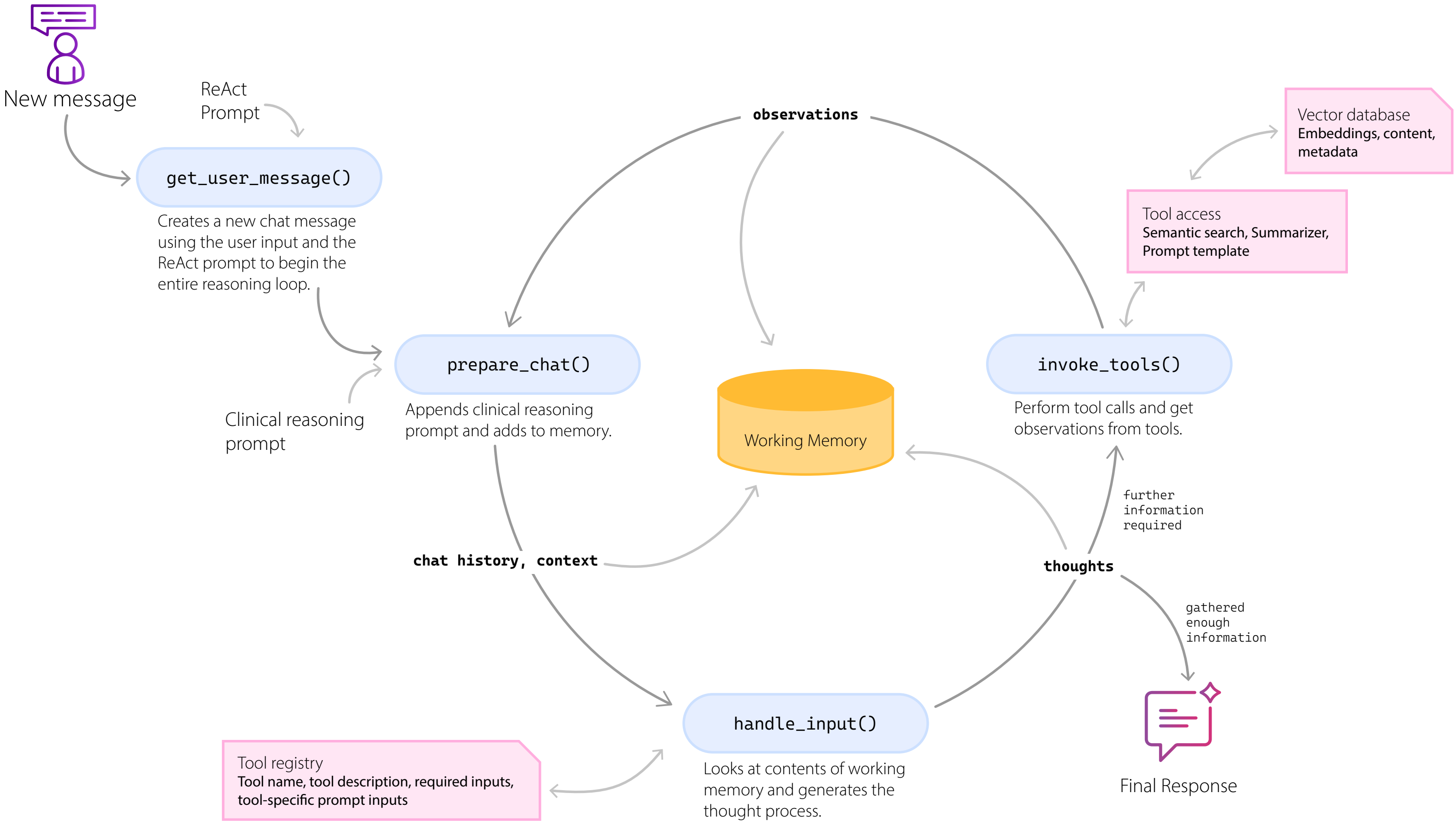


**Supplementary Figure 1**

A 33-year-old pregnant woman (24 weeks gestation) presents with metastatic HER2+ breast cancer with multiple liver metastases in September 2024. She is ER strongly positive, PR negative and HER2 positive (3+)
Given her pregnancy, how can the balance between effective cancer treatment and maternal-foetal safety be achieved?
What should be the next line of treatment?

*I need to look up guidelines on this matter. Looking up NCCN Guidelines for "pregnant woman with metastatic ER/PR negative HER2 positive breast cancer".*

**NCCN Guidelines:**
Treatment Guidelines for Pregnant Women with Metastatic HER2+ Breast Cancer

- General Principles
- First Trimester
- Second and Third Trimesters
- Monitoring
- Postpartum Management
- Insights

*I need some additional information to provide a more comprehensive answer tailored to this specific patient case. Looking up Drugs Information for "safety of trastuzumab during pregnancy".*

**Drugs:**
Trastuzumab and Safety During Pregnancy

- Mechanism of Action
- Pregnancy Considerations
- Indications
- Effects on Cancer Cells
- Safety Concerns
- Other Formulations

*The information on trastuzumab provides important insights into its mechanism of action and potential safety concerns during pregnancy. However, I still need more specific guidance on how to balance maternal and fetal risks in this patient case.*
*Looking up ASCO Guidelines for "treatment of HER2+ breast cancer during pregnancy".*

**ASCO Guidelines:**
Treatment guidelines for HER2+ breast cancer during pregnancy:

- Multidisciplinary team should be involved in managing pregnant patients undergoing neoadjuvant therapy.
- For HER2-positive disease in pregnant patients, the use of anti-HER2 therapy during pregnancy is not recommended due to potential risks to the fetus
- Post-delivery treatment follows standard guidelines for HER2+ breast cancer, emphasizing the importance of HER2-targeted therapies.
  - First-line treatment: Trastuzumab, pertuzumab, and taxane
  - Second-line treatment: Trastuzumab deruxtecan
  - Further lines: Other HER2-targeted combinations
- The high risk of recurrence in HER2+ disease underscores the need for close monitoring and prompt initiation of appropriate therapy after pregnancy.

### Balancing Effective Cancer Treatment and Maternal-Fetal Safety in Metastatic HER2+ Breast Cancer During Pregnancy

#### During Pregnancy
First Trimester: Systemic therapy is generally not recommended due to the high risk of fetal malformations.
Second and Third Trimesters: HER2-Targeted Therapy, Chemotherapy, Endocrine Therapy (Avoid until after delivery)

#### Postpartum Management
Resume standard HER2-targeted therapy

#### Monitoring
Foetal monitoring...
Consider early delivery...

#### Follow-Up
High risk of recurrence in HER2+ disease

#### Next Steps for This Patient
Engage in shared decision-making.
If treatment during pregnancy is desired, consider trastuzumab monotherapy with close fetal monitoring.
After delivery, promptly initiate standard first-line therapy

#### Key Insights
Delaying HER2-targeted therapy until after delivery is generally recommended

**Supplementary Figure 2**
